# Secondary attack rates of COVID-19 in Norwegian families: A nation-wide register-based study

**DOI:** 10.1101/2021.03.06.21252832

**Authors:** Kjetil Telle, Silje B. Jørgensen, Rannveig Hart, Margrethe Greve-Isdahl, Oliver Kacelnik

## Abstract

**Background:** Reported transmission rates of SARS-CoV-2 within families vary widely, and there are few reports on transmission from children to other family members. More knowledge is needed to guide infection control measures.

**Objective:** To characterize the family index case for detected SARS-CoV-2 and describe testing and secondary attack rates in the family.

**Design:** Register-based cohort study.

**Setting:** Individual-level administrative data of all families and all PCR tests for SARS-CoV-2 in Norway in 2020.

**Participants:** All families with at least one parent and one child below the age of 20, who lived at the same address (N=662 582), where at least one member tested positive for SARS-CoV-2 in 2020.

**Main outcome measures:** Secondary attack rates (SAR7) were defined as the share of non-index family members with a positive PCR test within seven days of the index case. SARs were calculated separately for parent- and child-index cases, and for parent- and child-secondary cases.

**Results:** We identified 7548 index cases, comprising 26 991 individuals, of which 12184 were parents and 14808 children. The index was a parent in 66% of the cases. Among the children, 42% of the index cases were in the age group 17-20 and only 8% 0-6 years. When the index was a parent, SAR7 was 24% (95%CI 24 to 25), whilst SAR7 was 14% (95%CI 13 to 15) when the index was a child. However, SAR7 was 24% (95%CI 20 to 28) when the index was a child aged 0-6 years and declined steeply with increasing age of the index child. SAR7 from index parent to other parents was 35% (95%CI 33 to 36), and from index child to other children 12% (95%CI 11 to 13). SAR7 from index child aged 0-6 to parents was 27% (95%CI 22 to 33). The percent of non-index family members tested within 7 days after the index case, increased from about 20% in April to 80% in December, however, SAR7 stabilized at about 20% from May.

**Conclusion:** Parents and older children are most often index cases for SARS-CoV-2 in families in Norway, while parents and young children more often transmit the virus within the families. This study suggests that whilst the absolute infection numbers are low for young children because of their low introduction rate, when infected, young children and parents transmit the virus to the same extent within the family.

| **What is already known on this topic** |
| --- |
| • Transmission of SARS-CoV-2 mainly occurs for prolonged and close contacts, in particular within families and households.<br>• How characteristics of the family index case and testing behaviour affect secondary attack rates (SAR) within families are largely unknown especially when young children are index cases. |
| **What this study adds** |
| • Nation-wide register data of all families in Norway suggest that children below school age are family index cases far less often than older children and parents, but secondary transmission within the family is just as likely when a child below school age is index as when a parent is index. Previous reports of low secondary attack rates in children within households may be due to few included children below school age. |

## Introduction

Despite a recent surge in studies of transmission of SARS-CoV-2 in families (Madewell *et al*.2020, Tian *et al*. 2020, ECDC 2020, Kuwelker *et al*. 2020), previously reported transmission rates vary widely. Estimates of transmission from children to other family members are particularly scarce (ECDC 2020, Grijalada *et al*. 2020) due to less testing and consequently low numbers of confirmed cases, and possibly more asymptomatic infections in children (Kim *et al*. 2020). One prospective study from the USA found secondary attack rates of over 50%, also from children below 12 years of age, but this study included just 5 index cases in this age group (Grijalada *et al*. 2020). Viner *et al*. (2021) conclude their systematic review by underlining the particular need for studies “that investigate secondary infections from child or adolescent index cases compared with secondary infections from adult index cases».

Understanding more about the roles of different types of index cases and transmission among family members is vital for containment strategies and contact tracing regimens. More reliable information on transmission from parents and children, both in and outside of the family, is important for decisions on family-wide quarantine measures, mitigation measures in schools and nurseries and limitations of extracurricular activities (Viner *et al*. 2021, ECDC 2020, Brandal *et al*. 2020). Ascertaining the role of index case characteristics in transmission of SARS-CoV-2 to other family members, has been difficult in previous studies because testing strategies have varied, and the number of families included has been limited. We provide a population-wide study of the testing and secondary infection rates for all affected families in Norway.

During 2020 about 1% (n=50 138) of the Norwegian population of 5.4 million was confirmed infected with SARS-CoV-2 by polymerase chain reaction (PCR) tests, and more than 3 million were tested (NIPH 2021). A trough in new infections in the summer, accompanied by many of the strict containment measures being loosened, was followed by a second peak in late autumn. During the first wave, testing capacity was limited, but there was still comprehensive testing of symptomatic patients and healthcare personnel. During the second wave, testing has been easily accessible and free for anyone with symptoms, as well as all close contacts of confirmed cases. All results from PCR tests for SARS-CoV-2 are transferred automatically and electronically to a national administrative system. This information can be combined with other information about individuals by merging registers based on unique personal identification numbers. These rich and population-wide data provide unique opportunities to improve our understanding of testing behavior and secondary attack rates (SAR) within all affected families by characteristics of family members.

The objective of this study was to describe characteristics of the index case and how testing behavior and SAR in the family varied with these characteristics.

## Methods

### Data

As part of the legally mandated responsibilities of The Norwegian Institute of Public Health (NIPH) during epidemics, a population-wide emergency preparedness register (*BeredtC19*) was established in cooperation with the Norwegian Directorate of Health (NIPH 2020). The purpose of the preparedness register is to provide a rapid overview and knowledge of how the pandemic and the measures that are implemented to contain the spread of the virus, affect the population’s health, use of healthcare services and health-related behaviors.

All laboratories in Norway conducting PCR tests for SARS-CoV-2 send information electronically to the Norwegian Surveillance System for Communicable Diseases (MSIS) when completing a test. Relying on the unique personal identification number provided to every resident in Norway at birth or upon immigration, test results can be linked at the individual level to demographic information in *BeredtC19* from the Norwegian Population Registry.

We utilized the individual-level data in *BeredtC19* for all residents of Norway, with vital demographic statistics (sex, year of birth, personal identification number of family members), and PCR test results for SARS-CoV-2 (test date, test result). Data on non-positive tests should be interpreted with caution prior to April 2020, as some laboratories did not report all non - positive tests before April. Institutional board review was conducted, and the Ethics Committee of South-East Norway confirmed (June 4th 2020, #153204) that an external ethical board review was not required.

### Population

Our study population included all members of families with at least one parent and one child aged 0-20 years residing on the same address on March 1^st^ 2020. We included families where at least one person was infected with SARS-CoV-2 according to a PCR test taken between March 1^st^ 2020 and January 1^st^ 2021. The data source did not contain non-residents (like tourists, temporary workers and asylum applicants).

### Definitions

Within each family, the index case was defined as the first family member who tested positive. Secondary cases included all non-index family members who tested positive by PCR within 7 days after the testing date of the index case (follow up period to January 31st 2021). We also recorded all non-index family members who were tested by PCR, regardless of test result, within 7 days after the testing date of the index case. Families where one unique index family member could not be identified, as more than one tested positive on the same date, were excluded.

In accordance with other studies, we calculated the proportion of secondary cases by the equation [number of family members with a positive test/number of family members] × 100, excluding the index case in both the numerator and denominator, and focusing on tests undertaken within seven days of the testing date of the index case (Grijalada *et al*. 2020, Park *et al*. 2020). We referred to this proportion as secondary attack rate within 7 days (SAR7). In a supplement, we also provided the proportion within 14 days (SAR14), as well as a plot of the proportion for each day up to 30 days after index testing date.

### Analyses

Descriptive statistics for the families were provided, including number of parents and children. The index case was described with respect to family position (parent, child, mother, father, son, daughter) and age (≤6, 7-12, 13-16, 17-20 years for children, and ≤30, 31-40,41-50, ≥50 years for parents). The overall percentage of non-index family members who were tested by seven days was calculated, and so was overall SAR7. Both the percentage tested and SAR7s were provided by characteristics of the index. We also calculated separately SAR7s from index child to parents, and from index parent to children. 95% confidence intervals (95% CI) around the SARs and percent tested were calculated using the Wilson method. The statistical software used was Stata MP v.16.

## Results

Among all Norwegian families with at least one parent and one child below the age of 20 and living at the same address as of March 1^st^ 2020 (N=662 582), we identified a total of 7 548 index cases with confirmed SARS-CoV-2 in 2020. The 7 548 families of the index cases comprised 26 991 individuals, of which 12 184 parents (45%) and 14 808 (55%) children (Table 1).

**Table 1.** Characteristics of family members of all families in Norway with at least one child and one parent, in which at least one family member tested positive for SARS-CoV-2 in a PCR test in 2020.

|  | All<br>(N=26 991) | Index<br>(N=7 548) | Non-index<br>(N=19 443) |
| --- | --- | --- | --- |
| Parents | 12 184 | 4 964 | 7 219 |
| Sex |  |  |  |
| ....Mother | 7 145 | 3 083 | 4 062 |
| ....Father | 5 038 | 1 881 | 3 157 |
| Age groups (years) |  |  |  |
| .... ≤30 | 747 | 458 | 289 |
| .... 31-40 | 3 612 | 1 767 | 1 845 |
| .... 41-50 | 5 221 | 1 894 | 3 327 |
| .... ≥50 | 2 603 | 845 | 1758 |
| Children | 14 808 | 2 584 | 12 224 |
| Sex |  |  |  |
| ....Daughter | 7 226 | 1 298 | 5 928 |
| ....Son | 7 582 | 1 286 | 6 296 |
| Age groups |  |  |  |
| ....≤6 | 3 631 | 200 | 3 431 |
| .... 7-12 | 4 352 | 517 | 3 835 |
| .... 13-16 | 3 575 | 781 | 2 794 |
| .... 17-20 | 3 250 | 1086 | 2 164 |
| Members in family |  |  |  |
| .... 2 | 2 854 | 1 427 | 1 427 |
| .... 3 | 7 047 | 2 349 | 4 698 |
| .... 4 | 9 412 | 2 353 | 7 059 |
| .... 5 | 5 256 | 1 043 | 4 213 |
| .... 6 or more | 2 422 | 376 | 2 046 |

Among the 7 548 index cases, there were 4 964 parents (66%) and 2 584 children (34%). There were more mothers (7 145) than fathers (5 038) in the families, and also among the index persons (3 038 vs 1 881). Among adults, the largest age group was 41-50 years, both in terms of total family members (5 221) and index persons (1 894). The youngest age group of adults (30 years and below) was the smallest with 747 persons, but had the largest proportion of index cases. Among the 14 808 children, the age groups were of relatively even size, but the number of index cases increased steeply with age, from 200 (8% of child index cases) among the youngest (aged 0-6) to 1 086 (42% of child index cases) among the oldest (aged 17-20).^1^

Both the proportion of tested family members and SAR increased steeply in the first days after the test date of the index case, and the additional increase was modest after 7 days and levelled off after 10 days (Supplement Figure A). The overall SAR7 was 21%, with 95% CI 20 to 21 (Table 2). When a parent was the index, SAR7 was 24% (95% CI 24 to 25), compared to 14% (95% CI 13 to 15) when a child was the index.

**Table 2.** Secondary PCR confirmed SARS-CoV-2 infections in non-index family members within seven days after index sampling date (SAR7). The data include all families in Norway consisting of at least one parent and one child, with at least one family member positive for SARS-CoV-2 in a PCR test in 2020. Percent (95% CI)

| Characteristics of the index case | Overall SAR7 | SAR7 among parents | SAR7 among children | Family members tested |
| --- | --- | --- | --- | --- |
| Overall | 21 (20-21) | 24 (23-25) | 19 (18-20) | 69 (69-70) |
| Parents | 24 (24-25) | 35 (33-36) | 21 (20-22) | 65 (64-66) |
| Sex |  |  |  |  |
| ....Mother | 23 (23-24) | 33 (30-35) | 21 (20-22) | 65 (63-66) |
| ....Father | 26 (25-27) | 37 (34-39) | 21 (20-22) | 65 (64-66) |
| Age groups (years) |  |  |  |  |
| .... ≤30 | 17 (15-20) | 34 (28-41) | 12 (10-15) | 56 (53-59) |
| .... 31-40 | 22 (20-23) | 32 (29-35) | 18 (17-20) | 62 (60-63) |
| .... 41-50 | 26 (25-28) | 36 (33-39) | 23 (22-24) | 69 (67-70) |
| .... ≥50 | 30 (28-32) | 37 (34-41) | 27 (24-29) | 66 (64-68) |
| Children (years) | 14 (13-15) | 15 (14-16) | 12 (11-13) | 78 (77-79) |
| Sex |  |  |  |  |
| ....Daughter | 13 (12-14) | 14 (12-15) | 11 (10-13) | 78 (77-79) |
| ....Son | 15 (14-16) | 17 (15-18) | 12 (10-14) | 78 (76-79) |
| Age groups |  |  |  |  |
| .... ≤6 | 24 (20-28) | 27 (22-33) | 19 (14-25) | 89 (86-92) |
| .... 7-12 | 14 (12-15) | 18 (15-20) | 9 (7-11) | 86 (84-87) |
| .... 13-16 | 14 (13-16) | 16 (14-18) | 12 (10-15) | 85 (83-86) |
| .... 17-20 | 11 (10-13) | 11 (10-13) | 12 (10-14) | 66 (65-68) |
| Members in family |  |  |  |  |
| .... 2 | 15 (13-17) | 12 (10-15) | 16 (14-19) | 64 (61-66) |
| .... 3 | 22 (20-23) | 25 (23-27) | 19 (17-20) | 68 (67-70) |
| .... 4 | 20 (19-21) | 24 (22-25) | 17 (16-18) | 71 (70-72) |
| .... 5 | 21 (19-22) | 25 (23-27) | 19 (17-20) | 71 (69-72) |
| .... 6 or more | 25 (23-27) | 27 (23-32) | 24 (22-27) | 66 (64-69) |

Note: Secondary attack rate (SAR7) was calculated as the number of non-index family members who tested positive within seven days after the date when the index family member tested positive, divided by all non-index family members. Percentage tested was calculated as the number of non-index family members who were tested within seven days after the date when the index family member tested positive, divided by all non-index family members. 95% CIs around the estimated secondary infection rates and percentage tested were calculated using the Wilson method.

SAR7 was 24% (95% CI 20 to 28) when the index was a child aged 0-6, but only 14% when the index was a child aged 7-12 or 13-16. The lowest SAR7 (11%, 95% CI 10 to 13) was found when a child aged 17-20 was the index. SAR7s were comparable for daughters (13%, 95% CI 12 to 14) and sons (15%, 95% CI 14 to 16) as index cases. SAR7s were slightly higher when the father was the index (26%, 95% CI 25 to 27) than when the mother was the index (23%, 95% CI 23 to 24).

SAR7 was 25% (95% CI 23 to 27) when there were six or more members in the family of the index case, and in the range 20-22 for families with 3-5 members. Families with two members had the lowest SAR at 15% (95% CI 13-17).

The highest SAR7 is found between adults (parents). SAR7 to parents was higher if a parent was the index case (35%, 95% CI 33 to 36) than if a child was the index case (15%, 95% CI 14 to 16) (Table 2, second column). For SAR7 to children, we saw the same tendency albeit weaker: SAR7 was 21% (95% CI 20 to 22) if a parent was the index case, and 12% (95% CI 11 to 13) if a child was the index case (Table 2, third column). There was more transmission to siblings if the index was in the youngest age group.

Testing rates within 7 days were lower when the index was a parent (65%, 95% CI 64 to 66) than when the index was a child (78%, 95% CI 77 to 79) (Table 2, fourth column). When the index case was a child, testing rates declined with the child’s increasing age, from 89% (95% CI 86 to 92) when the index was 0-6 years to 66% (95% CI 65 to 67) when the index was aged 17-20. When the index was a parent, testing rates were lowest when the parent was in one of the two lowest age groups (56% for ≤ 30, 62% for 31-40) and highest when the parent was in one of the oldest age group (78% for ≥50). The sex of the index person (for both children and adults) did not affect testing rates.

The percent of the non-index family members who were tested within seven days after the index case, grew from about 20% in April to about 80% in December (Figure 1). The associated SAR7 grew with the proportion of family members tested, up to when testing reached about 50% in May, after which the SAR7 remained relatively stable around 20% despite the testing proportion growing to 80%.

**Figure 1.**
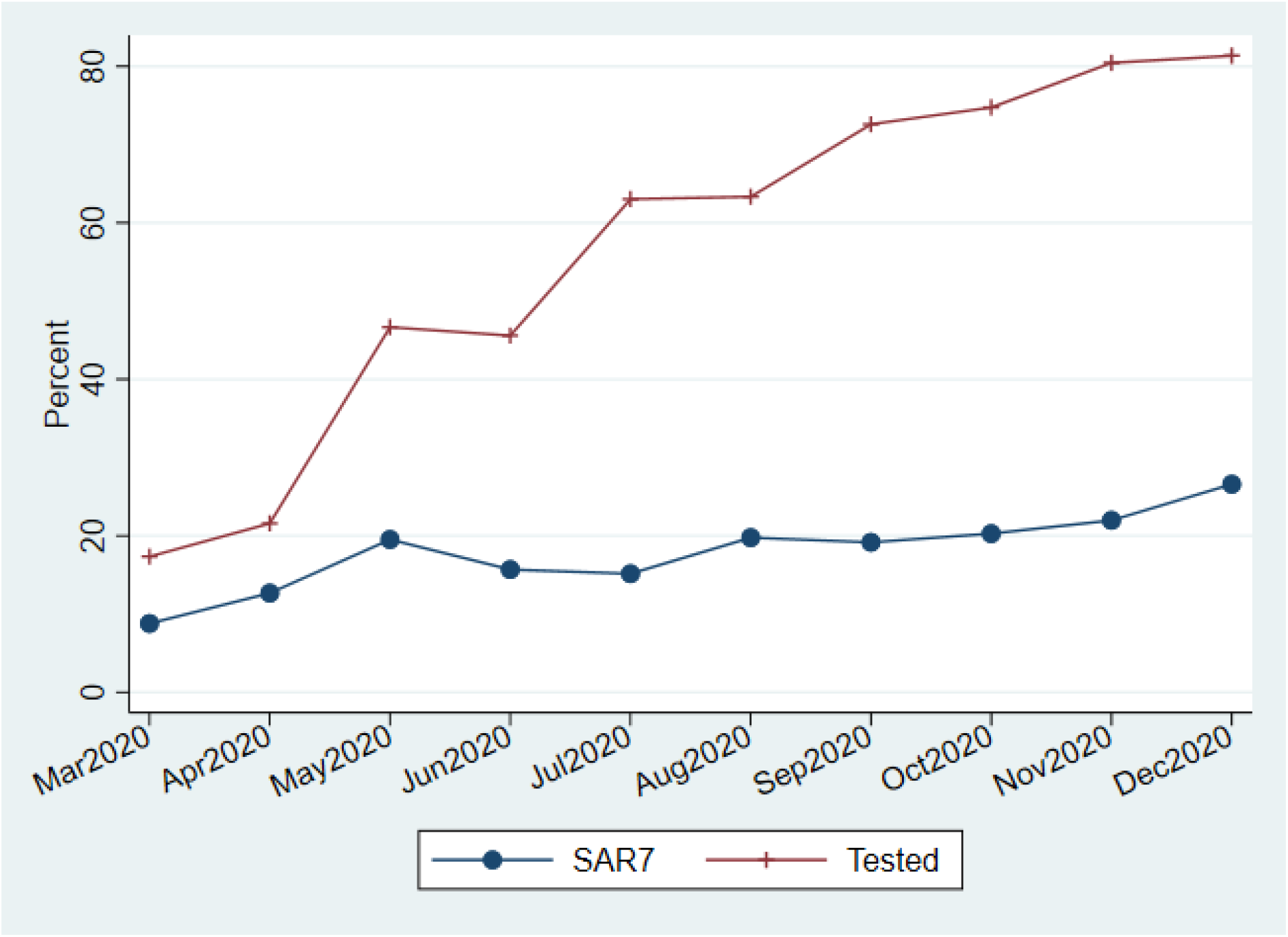
Monthly variation in **s**econdary PCR confirmed SARS-CoV-2 infections in non-index family members by 7 days after the date when index family member was positive (SAR7), and analogously for rate of non-index family members who were tested by 7 days (Tested). All families in Norway with at least one parent and one child, where at least one family member tested positive for SARS-CoV-2 in a PCR test in 2020.

## Discussion

In this nation-wide register study, we used SARS-CoV-2 PCR test results from all public and private Norwegian laboratories during 2020, combined with administrative population register data that identified all family members (parents and children) living at the same address.

### Principal findings

The index cases are mainly the parents (66%), even though there are fewer parents than children in the families. The children aged 17-20 years comprise 22% of all the children, but as many as 42% of the child index cases. The younger children are not often the index case. This could be because they display less symptoms, because it is easier to test an adult than a small child if both have symptoms, or because small children are less often infected outside of the family.

Subsequent transmissions within the families follow different patterns depending on the age of the index case. SAR7 was high both when a parent was the index (24%, 95% CI 24 to 25) and when a young child aged 0-6 was index (24%, 95% CI 20 to 28). However, it was low when an older child aged 17-20 was the index (11%, 95% CI 10 to 13). In short, parents and older children contribute the most to the introduction of SARS-CoV-2 into the family, while parents and young children contribute the most to transmitting the disease within the family. It could be that older children more often have another residence than the one they are registered at, i.e. in student accommodation or due to shared custody between parents. However, older children may also behave in ways that restrict viral transfer more than young children.

Parents transmit to other parents (35%, 95% CI 33 to 36) and less to children (21%, 95% CI 20 to 22), while children transmit similarly to parents (15%, 95%CI 14 to 16) and other children (12%, 95%CI 11 to 13).

Most cases of household transmission are detected by day seven after the index tested positive (SAR7 21%), increasing only somewhat to day 14 (SAR14 24%). In this study, the share of the non-index family members who had been tested was 69% (95%CI 69 to 70) by day 7, increasing to 72% (95% CI 72 to 73) by day 14.

The percent of the non-index family members who were tested within seven days was low in March and April (20%), but increased to 50% in May and to above 80% in December (Figure 1). While SAR7 grew in parallel with the percent tested from March to May, when testing rates grew beyond 50% from May, SAR7 remained around 20%.

### Comparison with related studies

Previous literature on intra-family transmission of SARS-CoV-2 is inconclusive, as the studies are few and small, with different designs, and report widely varying SARs (Madewell *et al*.2020, Tian *et al*. 2020, ECDC 2020, Grijalada *et al*. 2020, Kim *et al*. 2020, Park *et al*. 2020, Kuwelker *et al*. 2020, Viner *et al*. 2021).

We observed an overall SAR in line with previous studies, e.g. in two systematic reviews Madewell *et al*. (2020) reported household SAR of 17% and Lei *et al*. (2020) of 27%, both suggesting lower SARs to children than to adults. Viner *et al*. (2020) also estimate lower SAR from children than adults. However, comparison across studies is hard due to varying follow - up times, unclear handling of co-index cases, different testing regimes and small samples, especially for young child index cases. Very few studies calculate SARs across characteristics of the index or separately to parents and children. Grijalada *et al*. (2020) is a notable exception, but they only have five index children below the age of 12. Viner *et al*. (2020) find that their data were insufficient to conclude whether transmission of SARS-CoV-2 by children is lower than by adults, and they conclude their review by stating that studies “that investigate secondary infections from child or adolescent index cases compared with secondary infections from adult index cases are particularly needed».

### Interpretations

Norway has based much of its pandemic response on a demanding strategy of coordinated application of testing everyone with minor symptoms, isolation of positive cases, careful tracing of probable contacts and quarantine through the incubation period. The indications for testing, definitions of close contacts and length of quarantine have been regulated by law and adjusted over the course of the pandemic.

The observed increase in testing of family members reflects the increasing availability and reliance on testing throughout the pandemic. PCR-testing has been widely available in Norway, though in the first few months of the pandemic only symptomatic cases and healthcare personnel had wide access to the tests. From May and onwards, however, anyone wanting a test could get one by contacting their local municipal test-station, where testing was free of charge. This resulted in intensive testing as the infection rate started to grow from August, increasing the probability of persons with few or no symptoms to be included as index cases, and increasing the probability of secondary cases to be identified. It is worth noting that the frequency of testing for children, whilst generally lower than for the whole population, has been relatively high in relation to outbreaks in schools and nurseries.

Our study found that the recorded SAR7 was nearly as high as the SAR14. Almost all detected intra-family transmission occurs within the first seven days after the detection of an index case. This supports the current Norwegian strategy of testing and quarantine for 7-10 days for all family members after infection within the household.

An aggressive test/quarantine/isolate/-trace strategy can influence SAR and explain some of the variation in SAR in different studies. One would suspect that SAR7 is lower if the index case was tested in the presymptomatic period, as part of a contact tracing regimen, than if the index case was tested after symptom development, when the family would have been exposed for a longer time without prevention measures. We do not know how many of the index cases in our study who had symptoms, or whether they were tested because they were included in contact tracing around another case outside of the family.

We found substantial differences in the SAR depending on the characteristics of both the index case and the family composition. SAR was higher when young children (0-6 years) compared with older children were index, probably reflecting that the youngest need more close contact with their caregivers. Other studies (Madewell *et al*.2020, ECDC 2020, Kim *et al*. 2020, Park *et al*. 2020) have suggested that children have a lower attack rate and lower predisposition to serious disease and onward spread. After the lockdown period in March and April, when the nurseries and schools reopened, strict infection control measures were applied to prevent transmission in these institutions. To which extent these measures have been successful needs to be examined further, but our results underlines that children should be kept at home when they have symptoms that could indicate infection, and, moreover, that grandparents and other caregivers in risk groups for severe COVID-19 should not provide childcare for symptomatic children.

When the index case was a parent, there was a higher SAR towards the other parent than towards the children, and the SAR from young children was higher towards their parents than towards their siblings. The lower SAR rate we observe with increasing age of the index child most likely reflects the ability to identify cases earlier due to symptomatology, and also less close contact among the older children and the adults in the house. For very young children, it will likely be difficult to reduce contact even when contagion is detected. We also found a higher SAR associated with older parents than with younger ones. This may be due to a higher level of symptoms, and perhaps a higher level of caution resulting in more testing due to higher risk for serious disease among the oldest parents. However, we do not have data to explore this question.

As could be expected, a priori, we saw an increased SAR associated with the index living with a larger family. This reflects the larger number of contacts and probably more cramped living conditions. Norwegian advice has been that when isolating at home, cases should where possible, have their own bathroom, meals brought to them and as little contact with the rest of the house as possible. This is harder in situations of large families sharing smaller living spaces. Alternative housing has only been offered and accepted to a limited extent. Measures that make alternative housing more appealing, for instance moving the whole household to a larger dwelling rather than splitting out the index, may be considered.

### Potential limitations

The current Norwegian recommendations are that all close contacts should be tested at least once, preferably twice within a ten-day (recently seven day) period after the diagnosis of the index. In our study, the proportion of the non-index family members who had been tested by seven days after index date, is very high, but not 100%. It is thus possible that asymptomatic cases are sometimes not tested, and there is reason to assume that children below six years of age are overrepresented in this group, because they are more difficult to test. This may affect who we identify as index cases, and maybe also which secondary cases are identified. For example, when a young child (0-6) was index, about 90% of the family members were tested within seven days after index date, while this number was about 70% when a parent was index, which might suggest that more secondary cases were identified when index was a child than when index was a parent.

Better knowledge of actual directions of transmission within families requires prospective studies where all family members are tested daily with the same method in the week following index identification, preferably also with reporting of symptoms and genome identification of the viruses. Genome analysis would also help to reveal exposure to multiple infection events within the same family, which could interfere with the detected SAR. However, the incidence rate of SARS-CoV-2 has been low in Norway during the pandemic, estimated to peak at 74 per 100 000 per week toward the end of 2020 (NIPH 2021), which makes several transmission incidents into the same family at the same time less likely. A clear advantage of our study to such prospective studies, is that we do not have attrition: We observe every family, and we can observe all family members in the follow-up period, regardless of motivation to participate in a study or not. Indeed, our data stem from a real-world situation, where detection of secondary cases relates to a combination of the actual transmission of the virus and the behavioral responses to disease and the actual testing regime. This point is illustrated by us seeing lower SAR in the two first months of the pandemic, when testing capacity was limited, than later, when testing of family members was widely available. It seems that a test capacity where about 50% of the non-index family members are being tested, results in roughly the same SAR as a test regime where 80% of the non-index family members are tested. Another way of putting this, is that testing of cases with mild symptoms captures most of the cases. It should also be noted that we intentionally excluded families where there were “co-indexes” that could result in a greater infection pressure within the household.

Another limitation to our study is that the observation period does not include the coldest winter months, where people usually spend more time indoors, and when the climate is more favorable for viral sustainability on surfaces, and maybe also for transmission. Similar factors may also explain some of the variation in SARs between different studies. We observed a small increase in SAR during December, which may be a coincidence, or perhaps due to colder weather or closer contact between family members during the celebration of Christmas. Moreover, the introduction of variants of new and more easily transferable virus mutants could have played a role.

### Conclusions

By looking at register data for all families in Norway we see that parents and older children are most often index cases for SARS-CoV-2 detection. However, after introduction into the family unit, virus transmission within the family is more common from parents and preschool children than from older children. Detected infection rates among young children may be low, but this study suggests that infected young children transmit the virus within the family to the same extent as parents.

## Data Availability

The applied registry data contain sensitive information at the individual level and access are strictly regulated by Norwegian law.

https://www.fhi.no/sv/smittsomme-sykdommer/corona/norsk-beredskapsregister-for-covid-19/

## Acknowledgements

We would like to thank the Norwegian Directorate of Health, in particular Director for Health Registries Olav Isak Sjøflot and his department, for excellent cooperation in establishing the emergency preparedness register. We would also like to thank Gutorm Høgåsen and Anja Elsrud Schou Lindman for their invaluable efforts in the work on the register. We are grateful to Elisabeth Astrup, Ingeborg Elgersma, Tone B. Johansen and Karin Nygård for providing helpful comments to an earlier version of the manuscript. All interpretations and reporting of the data are the sole responsibility of the authors, and no endorsement by the register is intended or should be inferred. We would also like to thank everyone at the Norwegian Institute of Public Health who has been part of the outbreak investigation and response team.

## Conflict of interest disclosures

All authors have completed the ICMJE uniform disclosure form and declare: no support from any organization for the submitted work; no financial relationships with any organizations that might have an interest in the submitted work in the previous three years; no other relationships or activities that could appear to have influenced the submitted work.

## Funding/support

The study was funded by the Norwegian Institute of Public Health. No external funding was received.

## Role of the funder

The funding sources had no influence on the design or conduct of the study, the collection, management, analysis, or interpretation of the data, the preparation, review, or approval of the manuscript, or the decision to submit the manuscript for publication.

## Author contribution

Kjetil Telle designed the study and had access to all the data in the study and takes full responsibility for the integrity of the data and the accuracy of the statistical analysis. Oliver Kacelnik and Silje B. Jørgensen contributed to and critically evaluated the medical and contextual content of the analysis, and drafted the manuscript with Kjetil Telle. All authors contributed with conceptual design, analyses and interpretation of results. All authors contributed by critically revising the article for important intellectual content. All authors gave final approval for the version to be submitted.

## Funding/support

The study was funded by the Norwegian Institute of Public Health. No external funding was received.

## Supplement

**Supplement Figure A.**
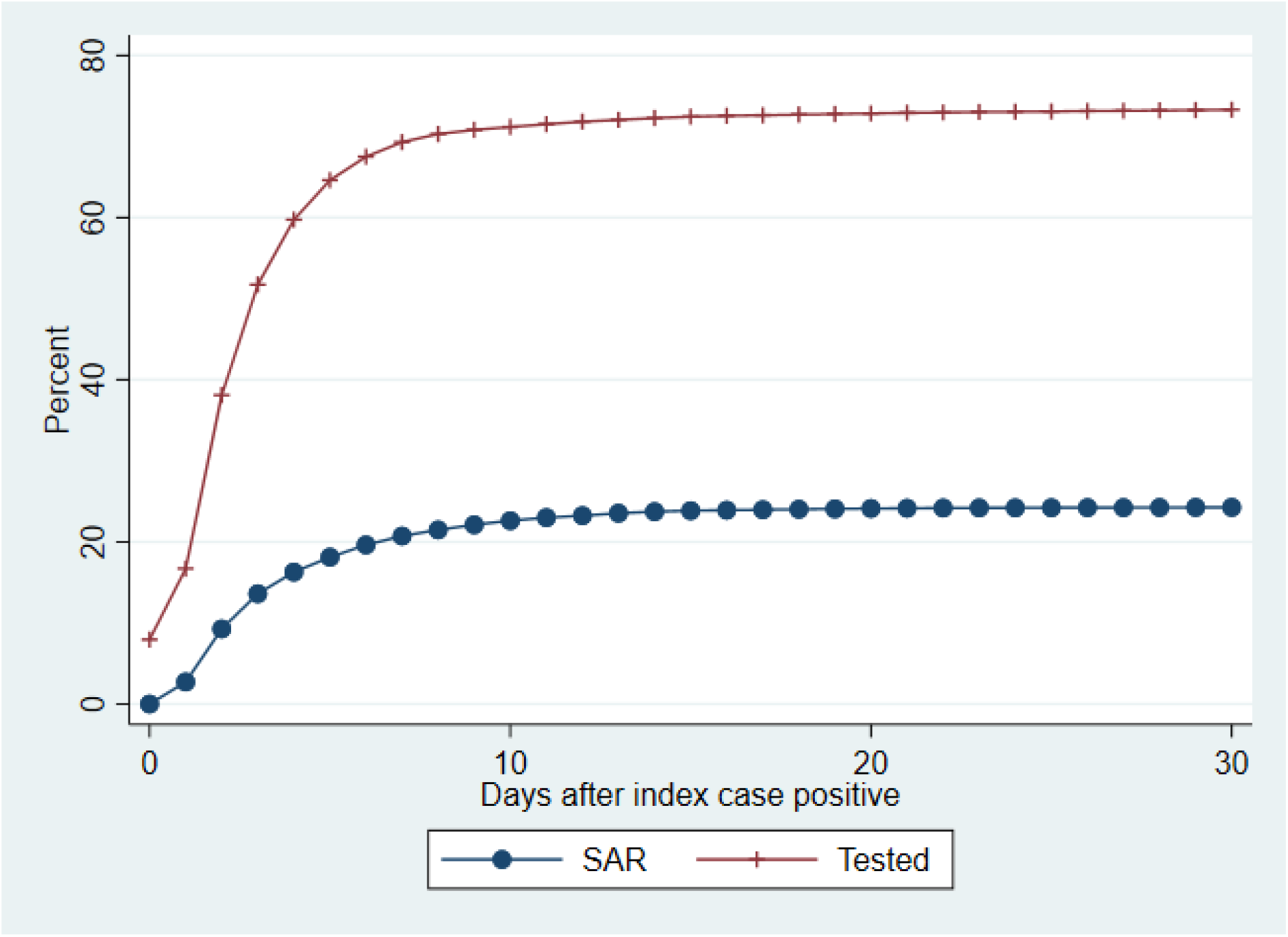
Secondary PCR confirmed SARS-CoV-2 infections in all non-index family members by given number of days after the date when index family member was positive, and analogously for percent of non-index family members who were tested. All families in Norway with at least one parent and one child, where at least one family member tested positive for SARS-CoV-2 in a PCR test in 2020.

**Supplement Table A.** Secondary PCR confirmed SARS-CoV-2 infections in non-index family members within 14 days after index sampling date (SAR14). The data include all families in Norway consisting of at least one parent and one child, with at least one family member positive for SARS-CoV-2 in a PCR test in 2020. Percent (95% CI)

| Characteristics of the index case | Overall SAR14 | SAR14 among parents | SAR14 among children | Family members tested |
| --- | --- | --- | --- | --- |
| Overall | 24 (23-24) | 27 (26-28) | 22 (21-23) | 72 (72-73) |
| Parents | 28 (27-28) | 38 (36-40) | 24 (23-25) | 68 (67-69) |
| Sex |  |  |  |  |
| ....Mother | 26 (25-27) | 36 (33-38) | 24 (23-25) | 68 (67-69) |
| ....Father | 29 (28-30) | 40 (38-42) | 25 (23-26) | 68 (67-69) |
| Age groups (years) |  |  |  |  |
| .... ≤30 | 30 (18-23) | 36 (30-43) | 16 (13-19) | 59 (55-62) |
| .... 31-40 | 24 (23-25) | 35 (33-38) | 21 (19-22) | 65 (64-66) |
| .... 41-50 | 30 (28-31) | 39 (36-42) | 26 (25-28) | 72 (70-73) |
| .... ≥50 | 34 (32-36) | 41 (37-45) | 31 (29-34) | 69 (67-71) |
| Children (years) | 17 (16-17) | 18 (17-19) | 15 (13-16) | 81 (80-82) |
| Sex |  |  |  |  |
| ....Daughter | 16 (15-17) | 16 (15-18) | 15 (13-17) | 80 (79-82) |
| ....Son | 17 (16-19) | 19 (18-21) | 15 (13-17) | 81 (80-82) |
| Age groups |  |  |  |  |
| ....≤6 | 27 (23-31) | 29 (24-34) | 23 (18-30) | 92 (89-94) |
| .... 7-12 | 17 (15-19) | 21 (19-24) | 12 (10-15) | 88 (86-89) |
| .... 13-16 | 17 (15-19) | 18 (16-21) | 15 (13-18) | 87 (86-89) |
| .... 17-20 | 14 (13-15) | 14 (13-16) | 14 (12-16) | 69 (68-71) |
| Members in family |  |  |  |  |
| .... 2 | 17 (15-19) | 13 (10-16) | 18 (16-21) | 67 (64-69) |
| .... 3 | 24 (23-25) | 28 (26-30) | 21 (20-23) | 71 (70-73) |
| .... 4 | 23 (22-24) | 27 (25-28) | 20 (19-21) | 74 (73-75) |
| .... 5 | 24 (23-25) | 28 (26-31) | 22 (21-24) | 73 (72-75) |
| .... 6 or more | 30 (27-32) | 32 (27-36) | 29 (26-32) | 71 (69-73) |

Note: Secondary attack rate (SAR14) was calculated as the number of non-index family members who tested positive within 14 days after the date when the index family member tested positive, divided by all non-index family members. Percentage tested was calculated as the number of non-index family members who were tested within 14 days after the date when the index family member tested positive, divided by all non-index family members. 95% CIs around the estimated secondary infection rates and percentage tested were calculated using the Wilson method.

## Footnotes

1 Except from an increase in adolescents among the index cases in the summer of 2020, we did not observe substantial changes in the age composition of index parents and children from May to December. Before May, the index cases were predominantly older adults (results not reported).

## Notes

### Competing Interest Statement

The authors have declared no competing interest.

### Funding Statement

The study was funded by the Norwegian Institute of Public Health. No external funding was received. The funding sources had no influence on the design or conduct of the study, the collection, management, analysis, or interpretation of the data, the preparation, review, or approval of the manuscript, or the decision to submit the manuscript for publication.

### Author Declarations

Institutional board review was conducted, and the Ethics Committee of South-East Norway confirmed (June 4th 2020, #153204) that an external ethical board review was not required. From our ethics comittee application: Knowledge of transmission, health service use and hospitalization for different geographic groups (e.g. municipality), in particular health personell, teachers and child care workers, waiters, taxi drviers etc is crucial for the emergency preparedness nationally and locally. Of particular importance are occupational risks for health personnel and others working in the health services, and how the authorities' measures to limit transmission (school lock downs, restrictions on travel, social distancing etc) impacts transmission to e.g. teachers, child care workers, hair dressers, taxi drviers etc. Further, it is important to study other sociodemographic factors of whom are being tested (age, sex, geography, etc.) and whom are testing positive (virus and antibodies), as well as how the testing and the results are within families.

